# Prediction of the clinical outcome of COVID-19 patients using T lymphocyte subsets with 340 cases from Wuhan, China: a retrospective cohort study and a web visualization tool

**DOI:** 10.1101/2020.04.06.20056127

**Authors:** Qibin Liu, Xuemin Fang, Shinichi Tokuno, Ungil Chung, Xianxiang Chen, Xiyong Dai, Xiaoyu Liu, Feng Xu, Bing Wang, Peng Peng

## Abstract

**Background:** Wuhan, China was the epicenter of the 2019 coronavirus outbreak. As a designated hospital, Wuhan Pulmonary Hospital has received over 700 COVID-19 patients. With the COVID-19 becoming a pandemic all over the world, we aim to share our epidemiological and clinical findings with the global community.

**Methods:** In this retrospective cohort study, we studied 340 confirmed COVID-19 patients from Wuhan Pulmonary Hospital, including 310 discharged cases and 30 death cases. We analyzed their demographic, epidemiological, clinical and laboratory data and implemented our findings into an interactive, free access web application.

**Findings:** Baseline T lymphocyte Subsets differed significantly between the discharged cases and the death cases in two-sample t-tests: Total T cells (p < 2·2e-16), Helper T cells (p < 2·2e-16), Suppressor T cells (p = 1·8-14), and TH/TS (Helper/Suppressor ratio, p = 0·0066). Multivariate logistic regression model with death or discharge as the outcome resulted in the following significant predictors: age (OR 1·05, p 0·04), underlying disease status (OR 3·42, p 0·02), Helper T cells on the log scale (OR 0·22, p 0·00), and TH/TS on the log scale (OR 4·80, p 0·00). The McFadden pseudo R-squared for the logistic regression model is 0·35, suggesting the model has a fair predictive power.

**Interpretation:** While age and underlying diseases are known risk factors for poor prognosis, patients with a less damaged immune system at the time of hospitalization had higher chance of recovery. Close monitoring of the T lymphocyte subsets might provide valuable information of the patient’s condition change during the treatment process. Our web visualization application can be used as a supplementary tool for the evaluation.

**Funding:** The authors report no funding.

## Introduction

The first case of COVID-19 infection was detected in December, 2019 in Wuhan, China. The situation quickly escalated into an epidemic all over China with tens of thousands of infections and several thousand deaths^1-2^. Further on March 11, 2020, the World Health Organization (WHO) declared COVID-19 a pandemic, pointing to the over 118,000 cases of infections in over 110 countries around the world and the sustained risk of further global spread^3-5^.

There is no known pre-existing immunity to COVID-19 in humans. In the absence of any anti-new corona virus therapy, infection of the virus weakens the host’s immune system, leading to Corona Virus Disease 2019 (COVID-19). Current estimates are that 2019-nCoV has an incubation period of 2 to 14 days, with potential asymptomatic transmission^6-7^.

Hubei province is located in the central region of China and Wuhan is the capital city of Hubei. With the traffic to and from the rest parts of the country, Wuhan had become the gateway for COVID-19 epidemic in China and is the most affected area in the country. The situation was further worsened by the Chinese Spring Festival travel rush (from mid-January to the end of February), when the world’s largest annual human migration occurs. On January 23, a lockdown in Wuhan and other cities in Hubei province started in an effort to quarantine the epicenter. Recent statistics suggest that almost 95% of reported COVID-19 cases in China originated from Wuhan ^2^.

Wuhan Pulmonary Hospital was a tertiary infectious disease hospital before the outbreak, with 610 beds and ten ICU beds. When the COVID-19 epidemic started in Wuhan, it became one of the first Covid-19 designated hospitals and has received more than 700 COVID-19 patients so far. In this study we aim to make full use of our experience with the COVID-19 patients and provide insight for the global community in evaluating the patient’s risk and progress. We have further implemented our findings into an interactive web tool for more practical uses.

## Methods

### Ethics approval

The study protocol was reviewed and approved by the Ethics committee of Wuhan Pulmonary Hospital (WPE 2020-8). Informed consents were obtained from all participants before enrollment. Patient records and information were de-identified prior to the analysis.

### Process Flow at the Hospital

Figure 1 illustrates the process flow in Wuhan Pulmonary Hospital. The patients were received from two sources: 1. Outpatient visit. People with symptoms such as fever, cough, or diarrhea made an Outpatient visit and took a blood test to exclude the possibility of bacterial infection. They then took an Influenza test to exclude the possibility of Influenza. Next a lung CT was taken to identified the infected area. Finally, a COVID-19 throat swab was taken to confirm the infection; 2. COVID-19 positive cases from the square cabin hospitals. These patients were transferred to Wuhan Pulmonary Hospital if their condition became severe or critical. Once the patient was hospitalized, regular examinations on T lymphocyte Subsets, COVID-19 PCR test, and lung CT were taken place to closely monitor the patient’s progress. Most patients who were hospitalized from January 28, 2020 to March 8, 2020 received treatment according to the fifth edition of the Ministry of Health guidelines^8-9^: 1. Bed rest and supportive treatment; 2. Regular examination on blood routine, CRP, biochemistry, coagulation function, myocardial enzymes, lung CT; 3. Oxygen therapy, including nasal catheter oxygen, transposal high-flow oxygen therapy, ventilator; 4. Antiviral therapy, including Lopinavir/Ritonavir tablets, Arbidol Hydrochloride Tablets, etc. If the patient maintained a normal body temperature for at least three days, and tested negative with COVID-19 (including throat swab, stool, and sputum) at least twice and 24+ hours apart, as well as showing significant improvement in lung CT, the patient would be considered recovered and discharged from the hospital. The discharged patients were transported by the Chinese government using designated vehicles and they were further isolated for at least two weeks.

**Figure 1:**
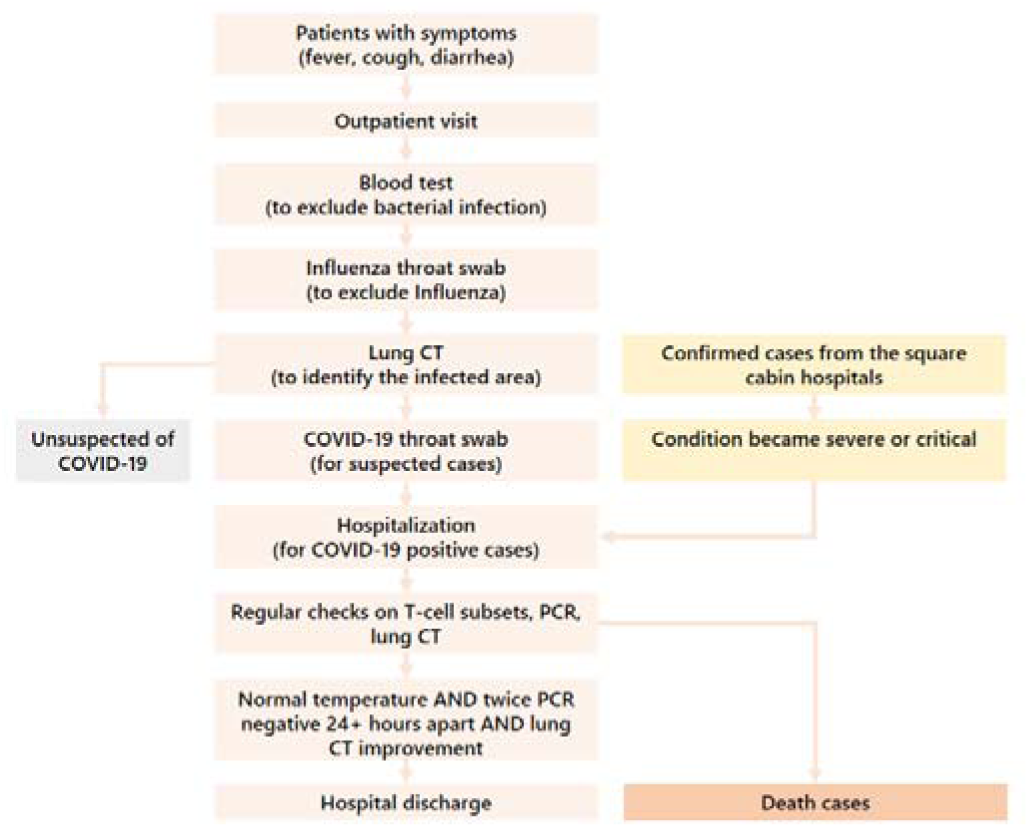
Process Flow at the Wuhan Pulmonary Hospital.

### Blood samples

2 mL venous blood samples were obtained from each patient to measure T cell subsets, and all the analyses were completed within 4 hours of sampling. FACSCalibur flow cytometer (BD Biosciences, San Jose, CA) was used for flow cytometry acquisition and analysis. The absolute count of each lymphocyte subset was determined using CD3/CD4/CD8/CD45 BD Multitest reagents according to the manufacturer’s protocol (BD Biosciences).

### Study population and data collection

From January 31 to March 8, 2020, a total of 721 patients who tested positive of COVID-19 were admitted to Wuhan Pulmonary Hospital. Among these patients, 430 completed the treatment and were discharged, 62 died, and 229 still remain in hospitalization. Excluding four patients whose direct cause of death was not COVID-19 infection, and selecting patients who had at least one T cell Subsets test available, we had a total of 340 patients in the study, including 310 discharged cases and 30 death cases. We reviewed laboratory test results and chest CT examinations of these 340 patients, and collected all the T lymphocyte subsets tests data. If multiple T lymphocyte subsets tests were performed, we chose the earliest one as the baseline. Two researchers independently reviewed the collected data to ensure data accuracy.

### Selection of analysis data

There have been a number of descriptive analyses about the epidemiological, clinical, laboratory, and radiological characteristics of the COVID-19 patients^10-14^. Little as yet known about how these characteristics can be used in guiding the practice of the healthcare providers^15^. As the world face the expanding COVID-19 pandemic, with the number of infected cases increase exponentially every day, any quick and easy method of understanding the patient’s condition can be valuable. With this in mind we seek to build a statistical model with only a few strong predictive characteristics. As several other research teams pointed out, the potential risk factors of older age, high SOFA score, and d-dimer greater than 1 µg/L could help clinicians to identify patients with poor prognosis at an early stage^12^. Older patients (>65 years) with comorbidities and ARDS are at increased risk of death^14^. Additionally, during our experience of treating the patients, we have noticed that the T lymphocyte subsets are closely correlated to the patient’s progress. We found that all the patients showed varying degrees of decline in T lymphocyte subsets at hospital admission. And the patient’s condition improved or worsened with the rise or fall of the T lymphocyte subsets. We have also considered some other immune indicators and inflammatory indicators, as well as the blood routine and lung CT, but they all have certain limitations. Immune indicators and inflammatory indicators such as thyrotropin and white blood cells were not as sensitive as the T lymphocyte subsets; the blood routine is not specific enough to differentiate the condition of the patients^11^, and although lung CT is an accurate way to assess the patient’s condition, it cannot be repeated too often especially for those critical patients who needed oxygen and ventilator. In conclusion, we decided to focus our research on patient’s age, underlying disease status, and the T lymphocyte subsets measures: Helper T cells, Suppressor T cells, and TH/TS (Helper T cells and Suppressor T cells ratio).

### Statistical analysis results and the interactive web tool

Among the 310 discharged cases, 155 were male and 155 were female. The average age was 56·4 years (SD 14·0). 107 (34·5%) of them had underlying diseases (the most common underlying disease is Hypertension, 23·9% for male and 20·0% for female). The average duration of the hospitalization was 11·1 days (SD 6·94), and it took 15·1 days (SD 10·3) on average for them to reach our hospital since their first symptoms (Table 1). Elderly patients tended to take longer to recover and they also had a higher probability to have underlying diseases (Figure 2a). The summary statistics for their baseline T lymphocyte subsets tests are the following: Total T cells percentage (mean 60·4%, SD 12·7%), Total T cells counts (mean 773, SD 549), Helper T cells percentage (37·2%, SD 10·6%), Helper T cells counts (mean 457, SD 342), Suppressor T cells percentage (mean 25·0%, SD 8·72%), Suppressor T cells counts (mean 297, SD 220), TH/TS (mean 1·71, SD 0·89), and Total Lymphocyte counts (mean 1160, SD 744) (Table 3).

**Table 1:**
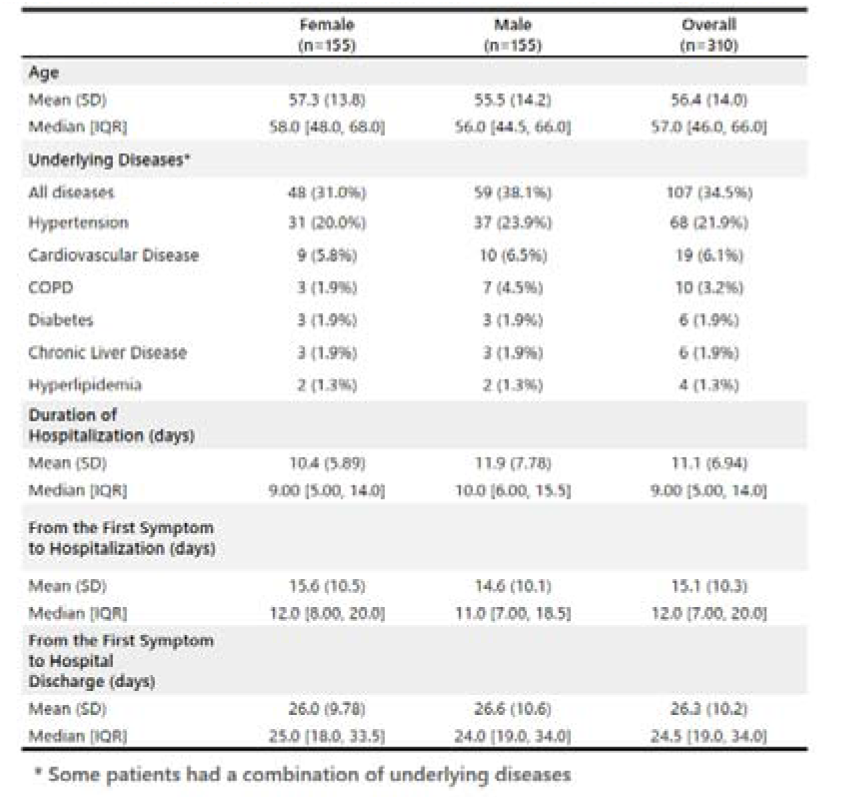
Summary table of the discharged patient characteristics and the duration of hospitalization

**Table 2:**
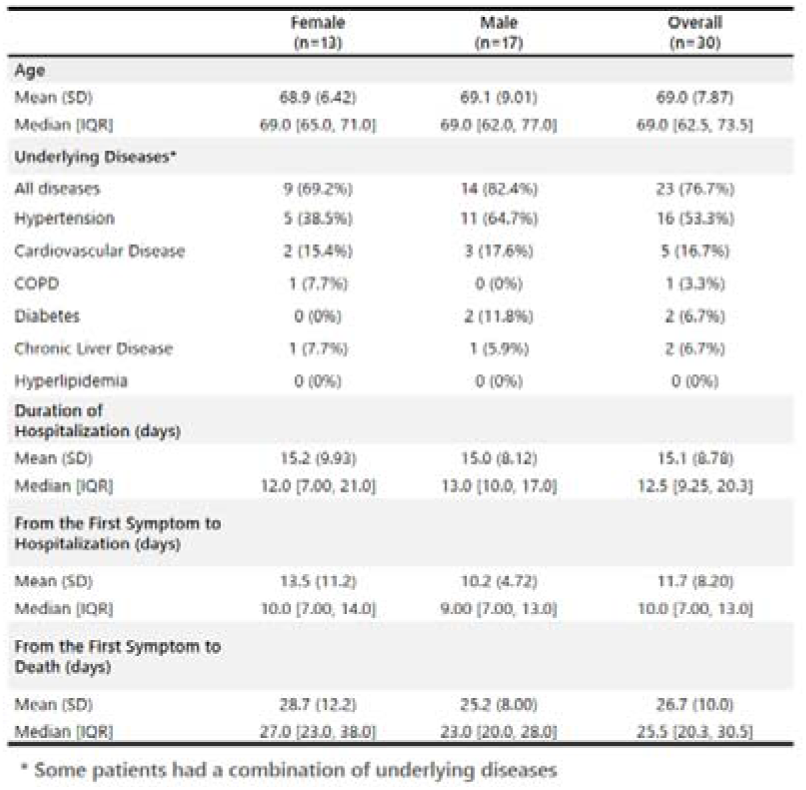
Summary table of the death patient characteristics and the duration of hospitalization

**Table 3:**
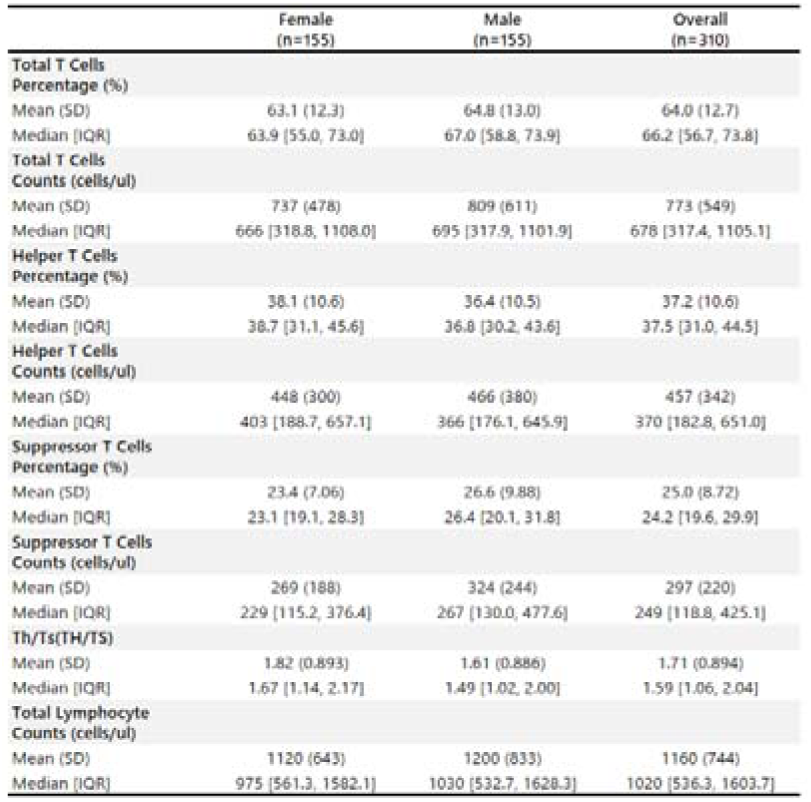
Summary table of the initial T cell test of the discharged patients

**Figure 2:**
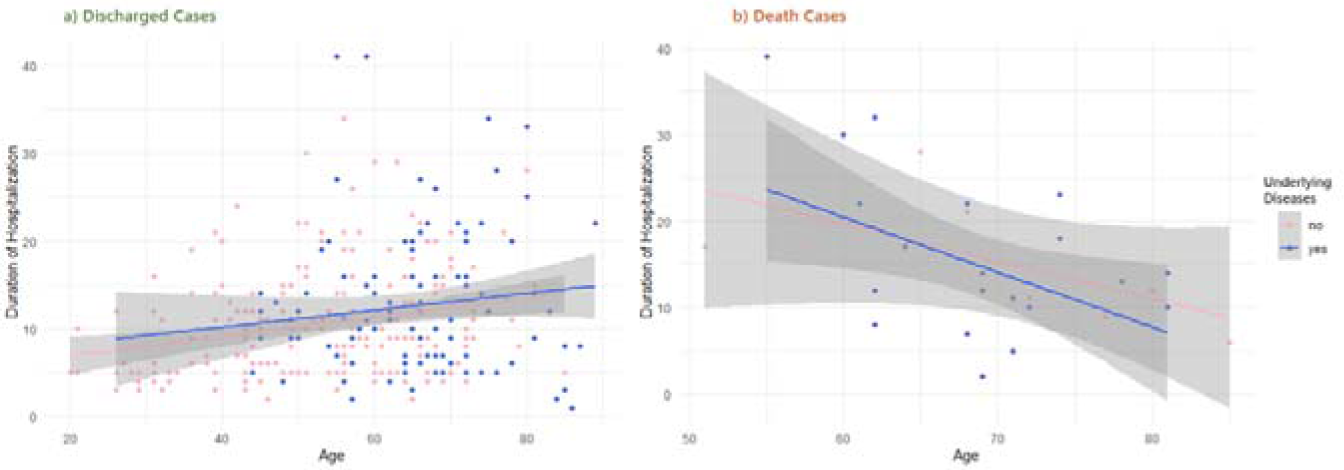
Duration of Hospitalization vs Age. a) Duration of the hospitalization plotted vs Age, for the discharged group. Pink dots indicate the patients with underlying medical conditions and the blue dots indicate those without. There are two observations from this chart: 1. Elderly patients tend to have underlying diseases than the younger ones; 2. The duration of the hospitalization tends to be longer among the elderly patients. The straight lines and the shaded areas show the regression line and the 95% confidence interval. b) Duration of the hospitalization plotted vs Age, for the death group. Pink dots indicate the patients with underlying medical conditions and the blue dots indicate those without. Elderly patients die more quickly than the younger ones. The straight lines and the shaded areas show the regression line and the 95% confidence interval.

Among the 30 death cases, 17 were male and 13 were female. The average age was 69·0 years (SD 7·87). 23 (76·7%) of them had underlying diseases (the most common underlying disease is also Hypertension, 64·7% for male and 38·5% for female). The average duration of the hospitalization was 15·1 days (SD 8·78), and it took 11·7 days (SD 8·20) on average for them to reach our hospital since their first symptoms (Table 2). The death event occurred more quickly among elderly patients (Figure 2b). The summary statistics for their baseline T lymphocyte subsets tests are the following: Total T cells percentage (mean 52·7%, SD 14·2%), Total T cells counts (mean 228, SD 168), Helper T cells percentage (33·8%, SD 11·8%), Helper T cells counts (mean 139, SD 98), Suppressor T cells percentage (mean 17·3%, SD 8·72%), Suppressor T cells counts (mean 80·9, SD 97·7), TH/TS (mean 2·41, SD 1·28), and Total Lymphocyte counts (mean 425, SD 254) (Table 4).

**Table 4:**
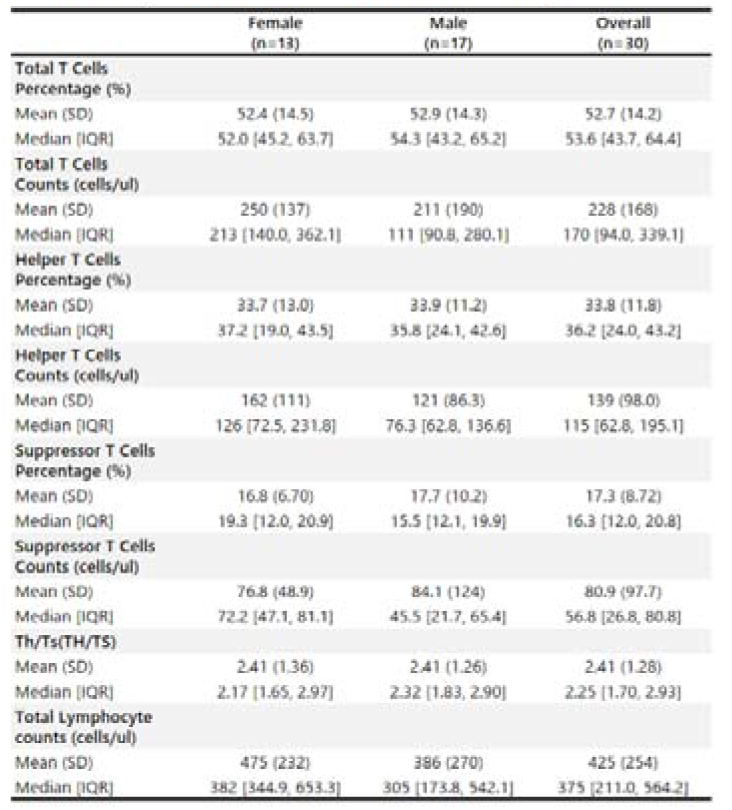
Summary table of the initial T cell test of the death patients

Two sample T-test between the discharged cases and the death cases in terms of their baseline T lymphocyte subsets measures yielded significant p values: Total T cells (p < 2·2e-16), Helper T cells (p < 2·2e-16), Suppressor T cells (p = 1·8e-14), and TH/TS (p = 0·0066) suggesting that patients with a poor prognosis had a more damaged immune system at the time of hospitalization (Figure 3).

**Figure 3:**
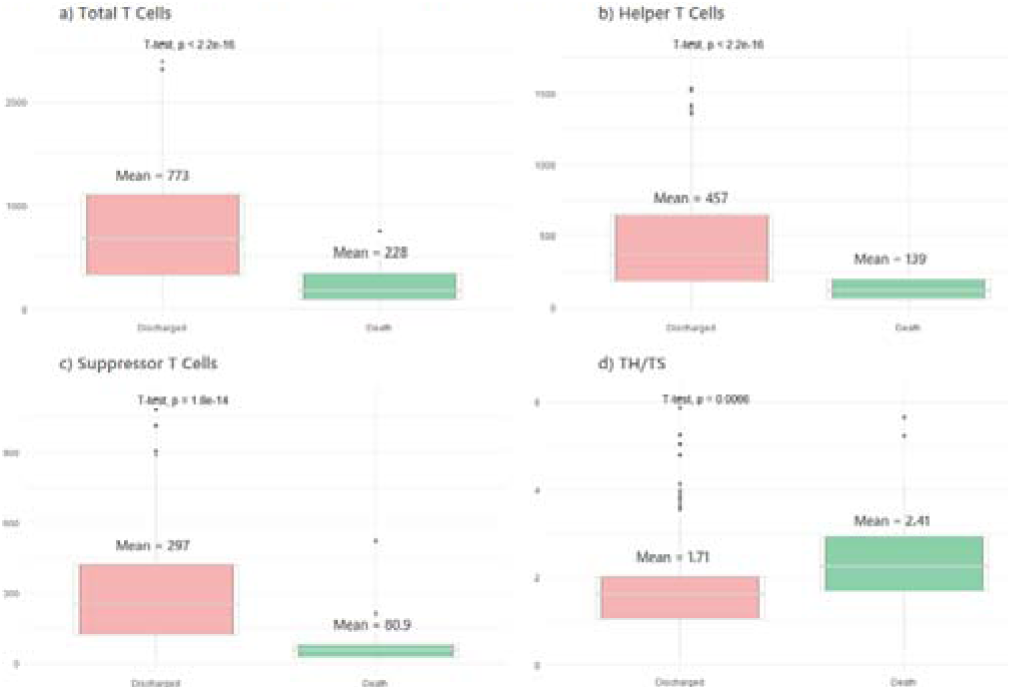
Comparison the T-cell Subsets between the Discharged Cases and the Death Cases. a) Boxplot of the difference in the total T cells between the last and the first T-cell Subsets tests during the hospitalization. T-test comparing the means of the two groups returns a p value less than 2.2e-16. b) Boxplot of the difference in the helper T cells between the last and the first T-cell Subsets tests during the hospitalization. T-test comparing the means of the two groups returns a p value less than 2.2e-16. c) Boxplot of the difference in the suppressor T cells between the last and the first T-cell Subsets tests during

Figure 4 are the scatterplots showing the pair-wise display of the three absolute T cell counts at the baseline: Helper T cells, Suppressor T cells, and the Total T cells (sum of the first two). Red dots indicate the death cases and green dots indicate the discharged cases. In all of the three scatterplots in Figure 4, there was a clear tendency that the death cases had lower cell counts than the discharged cases.

**Figure 4:**
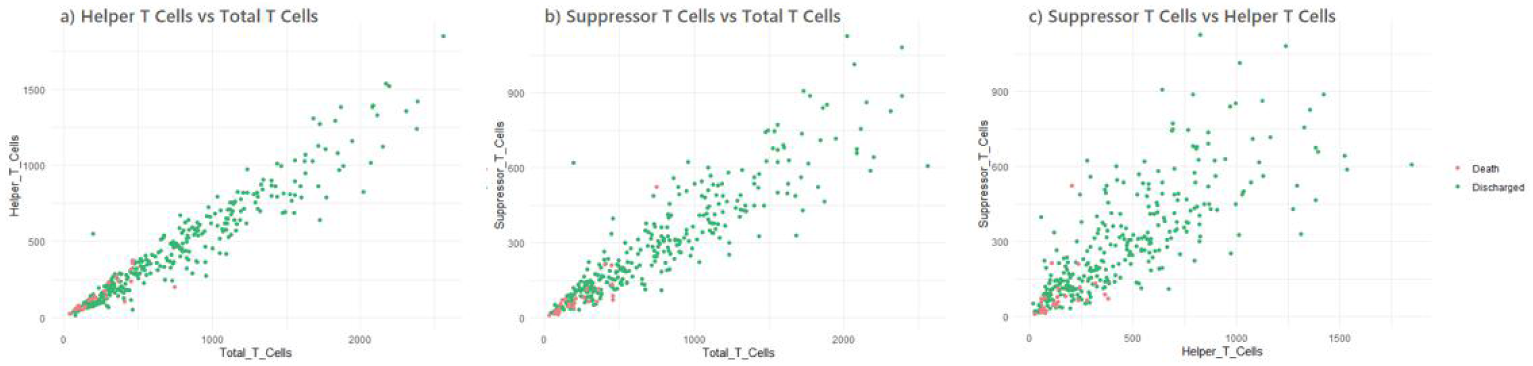
First Test Result after Hospitalization (Baseline) a) Helper T cells plotted vs total T cells, red dots indicate the death group and the green dots indicate the discharged group. The death cases have lower helper T cells and total T cells than the discharged group b) Suppressor T cells plotted vs total T cells, red dots indicate the death group and the green dots indicate the discharged group. The death cases have lower suppressor T cells and total T cells than the discharged group c) Suppressor T cells plotted vs helper T cells, red dots indicate the death group and the green dots indicate the discharged group. This is a different view from a) and b) showing the same tendency.

We also performed a multivariate logistic regression model using age, underlying disease status, and the baseline T lymphocyte subsets test as the predictors to predict the patient outcome (death or hospital discharge). The significant predictors are age (OR 1·05, p 0·04), underlying disease status (OR 3·42, p 0·02), Helper T cells on the log scale (OR 0·22, p 0·00), and TH/TS on the log scale (OR 4·80, p 0·00). The McFadden pseudo R-squared^16^ for the logistic regression model is 0·35, suggesting the model has a fair predictive power (The McFadden pseudo R-squared measure ranges from 0 to below 1, with values closer to 0 indicating lack of predictive power). Total T cells or Suppressor T cells, and Total Lymphocyte did not turn out to be significant predictors in our logistic regression model (Table 5).

**Table 5:**
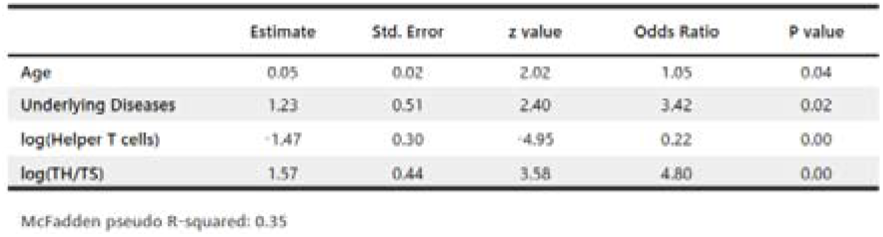
Multivariate Logistic Regression Model Result using Death as the Outcome

We believe by looking at some of the patient’s basic characteristics such as age and underlying diseases, together with the T lymphocyte subsets measures, could be a quick way to shed light on the patient’s prognosis, during the time of pressure and emergency. In order for our findings to be more applicable for public health workers fighting at the frontline, we have developed an interactive web data visualization tool to implement the algorithm and made it accessible for the world at the following web address: https://rpubs.com/mindyfang/covid19.

Simply by accessing to the web address and inputting a new patient’s information, the UI will show where the new patient is positioned (by a large blue dot) comparing to the reference panel of the 310 patients included in this research (Figure 5).

**Figure 5:**
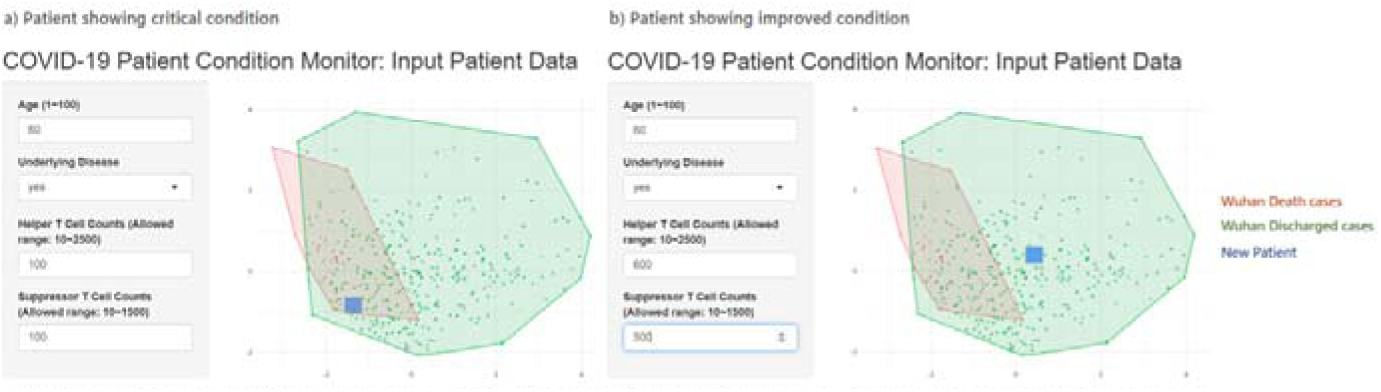
Screen Shots of the Interactive Web Tool. a) A 60 year old patient with underlying diseases, displayed by the large blue square, showing a critical condition with Helper T cell counts of 100 and Suppressor T cell counts of 100 b) The same patient showing improved condition, with Helper T cell counts increased to 600 and Suppressor T cell counts increased to 500

This data visualization tool applied a k-means clustering method to detect the differentiation in their baseline profile patterns among all patients. K-means clustering is a type of unsupervised learning method, enabling us to find and analyze the intrinsic underlying patient subgroups without any pre-defined subgroup labels. In real-world scenarios, clustering is a widely used technique for customer segmentation^17^.

The variables used in the k-means clustering included age, underlying disease status, Helper T cells (log scale), Suppressor T cells (log scale), and the Helper T cells and Suppressor T cells ratio (log scale) for the following reasons: 1. Age, underlying disease status, Helper T cells, and TH/TS were significant predictors in our multivariate logistic regression model. Although Suppressor T cells was not statistically significant, it was used in the TH/TS ratio. In fact a principal component analysis^18^ showed that all of the selected five indexes carry certain proportions of independent information (Figure 6a); 2. We did not use other lab tests such as the regular blood test items in our analysis because they are less differentiative than the T cell subset measures; and 3. We hope our interactive web tool could be utilized for quick use, therefore keeping as few input items as needed seems to be a more practical choice.

**Figure 6:**
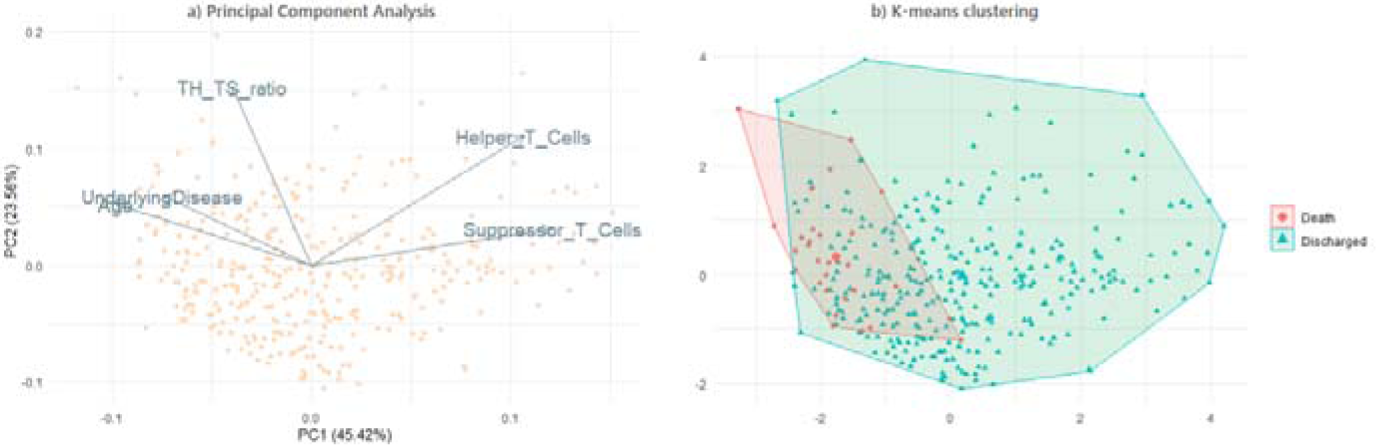
Principal Component Analysis of the T-cell subsets. a) The three selected T-cell subsets indexes and age, underlying disease indicator were plotted against their first two principal components, which carries 45.42% and 23.56% of the total information respectively. The dots in the background represent each patients. There was very little overlap among the six variables suggesting them carrying exclusive information. b) Multi-dimentional transformation of the above five variables for the 340 patients in our study, so that the underlying patterns can be recognized to the maximum extent. A proportion of the discharged cases had similar T-cell subsets profiles with the death cases which made it difficult to differentiate between them.

Figure 6b shows the k-means clustering result using the Wuhan Pulmonary Hospital data. After multi-dimensional data transformation, the algorithm separates the death group and the discharged group as shown in the graph. A proportion of the discharged cases had similar profiles with the death cases, which had made it difficult for the algorithm to differentiate them apart. However, by using this algorithm, it is possible to identify a large number of patients with relatively good prognosis.

We have also uploaded a dummy date set with de-identified and randomly modified patient data, as well as all the source code used for the current analysis as well as the interactive web application to: https://github.com/mindy-fang/COVID-19. All of our source code were written with the R programming language and the interactive web application was developed with the shiny package^19-21^ (Rstudio Version 1.2.1335, R version 3.6.0). Other fellow researchers can substitute the dummy data with their own data, or modify the source code to make their own applications. Meanwhile we will keep updating our reference panel as we include more patients, so that the algorithm would gain more and more statistical power over time.

## Discussion

To our best knowledge, the current research is the largest retrospective study of COVID-19 patients with known clinical outcomes so far. Significant reductions in T cells are very common in severe COVID-19 patients. Age-dependent deficits in T cell and B cell functions and overproduction of type 2 cytokines may cause inadequate viral replication control and longer pro-inflammatory responses, which may lead to poor prognoses^22^. Lymphopenia is a prominent part of SARS-CoV infection and lymphocyte counts may be useful in predicting the severity and clinical outcomes^23-25^. We know that 2019-nCoV was once called Severe Acute Respiratory Syndrome Coronavirus 2 (SARS-CoV-20)^26-27^. The level and the speed of T cell recovery are important factors of assessing disease prognoses and guiding early intervention in critically ill patients. In this study we have identified that older age, underlying diseases, and low T cell counts may be risk factors for poor clinical outcomes in COVID-10 positive patients. We have also implemented an interactive web tool to visualize a new patient’s risk using these risk factors. When a new patient’s data is entered through the web UI, it will be shown where he is as compared to the 310 patients included in this study.

It should be noted that our intension is not for the web tool to be considered as a 100% gold standard for determining the final clinical outcome of the patients. The visualization result simply suggests whether the new patient is more likely to recover or to have adverse outcomes. Another important significance of our tool is the monitoring of patient’s progress in real time during the treatment. For example, the patient’s longitudinal profile gradually shifting towards the red centroid (poor outcome) or the green centroid (good outcome) may provide insight on the patient’s progress (Figure 5). However, the actual final prognosis of the patient depends on many other factors, including the starting time of the treatment, treatment compliance, degree of treatment, and so on.

Our algorithm has the following limitations: It may not predict accurately for younger patients, or patients with no symptoms, since our training data contains relatively old and severe patients; Secondly, the model was built based on a sample size of 340, which is not a large number. But we will keep updating our web application as we collect more data.

In addition, the reference panel used in our analysis were infected population in Wuhan, China. Although there was no clear evidence that the underlying mechanism of the T cell depletion under the COVID-19 infection is similar or different across ethnicity groups, it is only natural to assume that the baseline values and the degree of T cell depletion would be slightly different. We found that the T-lymphocytes, B-lymphocytes, and NK cells did differ among people in different regions^28-30^. We encourage researchers around the world to download the source code and customize it with their own data, as they accumulate more experience and knowledge with patients from their own hospitals or regions.

To end our writing with the most recent update (March 13, 2020), the epidemic in Wuhan has gradually passed its peak. All the cabin hospitals were closed, and other hospitals in Wuhan started to return to their normal track. Medical volunteer teams have returned to their home cities. We give thanks to all the support that we have received during our most difficult time and hope the situation would improve quickly for other parts of the world.

## Data Availability

The data used to support the findings of this study are available from the corresponding author upon request.

## Acknowledgments

Our research was supported by medical teams from Shanxi, Inner Mongolia and other parts of China. The authors would like to thank Prof Li Ming, Prof Zhu Qi, Prof Yang Chengqing, Prof Guo Guangyun, Prof Du Juan, Prof Du Ronghui for their technical support and Prof Chen Xianxiang for guidance in interpretation of the results.

## Notes

### Competing Interest Statement

The authors have declared no competing interest.

